# Pharmacometabolomics Identifies Candidate Predictor Metabolites of an L-carnitine Treatment Mortality Benefit in Septic Shock

**DOI:** 10.1101/2021.01.28.21250687

**Authors:** Michael A. Puskarich, Theodore S. Jennaro, Christopher E. Gillies, Charles R. Evans, Alla Karnovsky, Cora E. McHugh, Thomas L. Flott, Alan E. Jones, Kathleen A. Stringer, On behalf of the RACE Trial Investigators

**Affiliations:** Department of Emergency Medicine, University of Minnesota, 420 Delaware Street SE, Minneapolis, MN USA 55455; Department of Emergency Medicine, Hennepin County Medical Center, 420 Delaware Street SE, Minneapolis, MN USA 55455; The NMR Metabolomics Laboratory and the Department of Clinical Pharmacy, College of Pharmacy, University of Michigan, 428 Church Street, Ann Arbor, MI USA 48109-1065; Department of Emergency Medicine, University of Mississippi Medical Center 2500 N State St, Jackson, MS USA 39216; Michigan Center for Integrative Research in Critical Care (MCIRCC), University of Mississippi Medical Center 2500 N State St, Jackson, MS USA 39216; Michigan Institute for Data Science, Office of Research; University of Mississippi Medical Center 2500 N State St, Jackson, MS USA 39216; Michigan Regional Comprehensive Metabolomics Resource Core ((MRC)^2^), University of Mississippi Medical Center 2500 N State St, Jackson, MS USA 39216; Division of Metabolism, Endocrinology and Diabetes, Department of Internal Medicine, University of Mississippi Medical Center 2500 N State St, Jackson, MS USA 39216; Department of Computational Medicine and Bioinformatics, University of Michigan School of Medicine; University of Mississippi Medical Center 2500 N State St, Jackson, MS USA 39216; Division of Pulmonary and Critical Care Medicine, Department of Internal Medicine, School of Medicine, University of Michigan, 7300 Medical Science Building I, 1301 Catherine Street, Ann Arbor, MI USA 48109-5624

**Keywords:** sepsis, metabolomics, acylcarnitines, branched chain amino acids, nuclear magnetic resonance, liquid chromatography-mass spectroscopy

## Abstract

**Background:** Sepsis-induced metabolic dysfunction contributes to organ failure and death. L-carnitine has shown promise for septic shock, but a recent study demonstrated a non-significant reduction in mortality.

**Methods:** A pharmacometabolomics study of patients (n=250) in a Phase II trial of L-carnitine to identify metabolic profiles predictive of a 90-day mortality benefit from L-carnitine. The independent predictive value of each pre-treatment metabolite concentration, adjusted for L-carnitine dose, on 90-day mortality was determined by logistic regression. A grid-search analysis maximizing the Z-statistic from a binomial proportion test identified specific metabolite threshold levels that discriminated L-carnitine responsive patients. Threshold concentrations were further assessed by hazard-ratio and Kaplan-Meier estimate.

**Findings:** Accounting for L-carnitine treatment and dose, 11 ^1^H-NMR metabolites and 12 acylcarnitines were independent predictors of 90-day mortality. Based on the grid-search analysis numerous acylcarnitines and valine were identified as candidate metabolites of drug response. Acetylcarnitine emerged as highly viable for the prediction of an L-carnitine mortality benefit due to its abundance and biological relevance. Using its most statistically significant threshold concentration, patients with acetylcarnitine ≥35µM were less likely to die at 90 days if treated with L-carnitine (18 g) versus placebo (p=0.01 by log rank test).

**Interpretation:** Metabolomics identified independent predictors of 90-day sepsis mortality. Our proof-of-concept approach shows how pharmacometabolomics may be useful for tackling the heterogeneity of sepsis and informing clinical trial design. Also, metabolomics can help understand mechanisms of sepsis heterogeneity and variable drug response, since sepsis induces alterations in numerous metabolite concentrations.

## Introduction

Sepsis represents the leading cause of death in the intensive care unit and the single most expensive inpatient diagnosis, representing more than $17 billion in healthcare costs annually in the United States.^1-3^ Septic shock carries a particularly poor prognosis, with short-term mortality rates of approximately 40%. Among the many physiologic disturbances associated with sepsis is a profound shift in metabolism.^4^ Hyperlactatemia represents one of the hallmarks of sepsis and is now considered a criterion for the diagnosis of septic shock.^5^ However, hyperglycemia, lipolysis, and protein catabolism are also common and similarly associated with increased mortality.^4,6^ Manipulation of these processes represents an underdeveloped but promising target for novel pharmacotherapies.

Despite the concerning sepsis mortality statistics and an increasingly focused research effort on the condition, clinical trials of novel sepsis pharmacotherapies have traditionally yielded disappointing results. While the causes of the failure of clinical trials to further novel treatments are multifactorial, the highly heterogeneous nature of sepsis certainly contributes to these results.^7,8^ This highlights the need to forge a better understanding of the heterogeneity and complexity of the clinical illness by identifying sepsis endotypes.^9^ In doing so, strategies for enriched patient selection could be used to improve the precision of clinical trials. Importantly, predictive and prognostic enrichment strategies for clinical trials have been advocated by many and have been issued as guidance by regulatory agencies like the U.S. Food and Drug Administration.^10-12^

We recently completed a phase II, Bayesian adaptive dose-finding randomized control trial comparing L-carnitine (6, 12, or 18 g) treatment to saline (placebo) for the early treatment of septic shock. None of the tested doses of L-carnitine resulted in a significant reduction in sequential organ failure assessment (SOFA) score at 48 hours, though the highest and best performing dose (18 g) demonstrated a non-significant 3% and 6% absolute mortality reduction at 28 days in the intention to treat and per protocol analyses compared to saline placebo, respectively.

In parallel with the planning of the original trial, we designed an ancillary metabolomics study, the L-Carnitine Pharmacometabolomics in Sepsis (CaPS) study, to identify candidate metabolites of drug response that could serve to endotype a heterogeneous septic shock cohort and direct the design of a clinical enrichment strategy for a phase III trial. A number of studies have demonstrated the importance of energy-related metabolites for the differentiation of sepsis survivors and the identification of sepsis endotypes,^4,6,13-16^ most of which are readily detected by nuclear magnetic resonance (NMR) spectroscopy^6,14,15^ and targeted liquid chromatography – mass spectroscopy (LC-MS) assays.^16^ Furthermore, we have previously demonstrated the utility of metabolomics in predicting drug response (pharmacometabolomics) in sepsis^15^ employing relatively quantified NMR metabolites and acylcarnitines generated by an LC-MS assay. With this background in mind, we hypothesized that serum concentrations of acylcarnitines and/or other metabolites could differentiate patients that disproportionately benefit from L-carnitine treatment as measured by mortality.

## Methods

### Study design

This study utilized pre-treatment serum samples collected from 236 of the 250 patients enrolled in the Rapid Administration of Carnitine (RACE) in Sepsis clinical trial.^17^ The parent trial was approved by each site’s institutional review board, all patients or their surrogate gave written informed consent, and it was registered at clinicaltrials.gov prior to initiation (NCT 01665092). Details of the blood samples included in the study are provided in the supplementary material, Figure 1. Serum samples were assayed for acylcarnitines by LC-MS^16^ and by quantitative proton (^1^H) nuclear magnetic resonance (NMR) as previously described.^18,19^ More details about the methods for these measurements can be found in the supplementary material.

**Figure 1:**
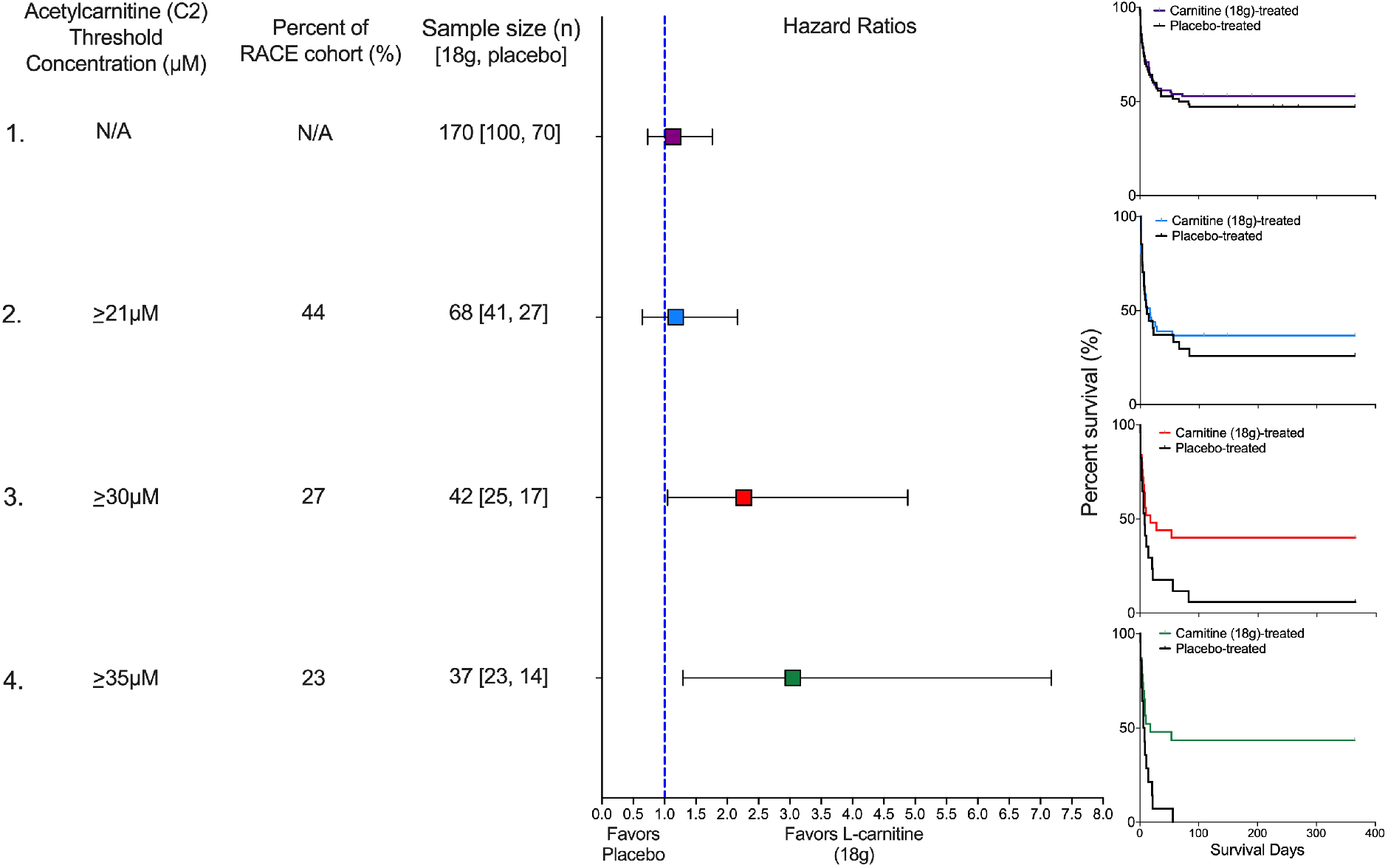
Pre-treatment acetylcarnitine (C2) concentration as a predictive clinical trial enrichment strategy. We present four scenarios to illustrate how different threshold concentrations of acetylcarnitine (C2), a high abundant acylcarnitine, would have impacted the outcome of the Rapid Administration of Carnitine (RACE) in Sepsis clinical trial in patients treated with either L-carnitine (18 g) or placebo. In scenario 1, no threshold concentration is used so the entire RACE cohort (n=236) is eligible. The sample size of 170 patients represents those that received either L-carnitine (18 g; n=100) or placebo (n=70). The hazard ratio is not significant, and consistent with the parent trial, the Kaplan-Meier curve shows no mortality benefit of L-carnitine (p=0·57). In scenario 2, an acetylcarnitine (C2) threshold concentration of ≥21µM is used. Forty-four percent (n=104) of the RACE cohort met this criterion and of these, 68 patients received either L-carnitine (18 g) or placebo. The hazard ratio is not improved, and the Kaplan-Meier curve shows no mortality benefit of L-carnitine (p=0·59). In scenario 3, an acetylcarnitine (C2) threshold concentration of ≥30µM is used. Twenty-seven percent (n=64) of the RACE cohort met this criterion and of these, 42 patients received either L-carnitine (18 g) or placebo. The hazard ratio is significant and favors L-carnitine (18 g); the Kaplan-Meier curve shows a mortality benefit of L-carnitine (p=0·04). Finally, scenario 4 uses the acetylcarnitine (C2) concentration associated with the maximum Z-statistic (see Table 4 in the supplementary material), ≥35µM. Twenty-three percent (n=54) of the RACE cohort met this criterion and of these, 37 patients received either L-carnitine (18 g) or placebo. The hazard ratio is significant, and the Kaplan-Meier curve shows a mortality benefit of L-carnitine (p=0·01). Kaplan-Meier curves were interpreted using log-rank (Mantel-Cox) tests; hazard ratios were calculated using Mantel-Haenszel.

### Outcomes

We elected to use mortality as the outcome of our analysis because the primary end point of the RACE trial (reduction in SOFA score at 48 h) was not met, but the 18 g dose of L-carnitine resulted in a trend towards a reduction in mortality. Based on data suggesting a substantial continued decline in mortality among sepsis patients beyond 28 days and preliminary data from our phase I data suggesting continued benefit from L-carnitine treatment on longer term mortality rates,^20^ we elected to assess the cumulative distribution mortality function to find the optimal time frame for assessment of mortality (28, 90, 180, or 365 days). By 90-days, ∼90% of the deaths had occurred, (see Figure 2 in the supplementary material), based on this analysis, we chose 90-day mortality as the primary clinical outcome.

**Table 1:** Demographics and clinical characteristics of the cohort, stratified by 90-day mortality

| Variable | Survived (n = 111) | Died (n = 125) | p value |
| --- | --- | --- | --- |
| Demographics |  |  |  |
| Age, years (IQR) | 61 (49, 69) | 66 (57, 76) | 0.002 |
| Male, n (%) | 60 (54) | 74 (41) | 0.43 |
| Female, n (%) | 51 (46) | 51 (59) |  |
| Race |  |  |  |
| Black, n (%) | 33 (30) | 39 (31) | 0.88 |
| Asian, n (%) | 3 (3) | 2 (2) |  |
| White, n (%) | 68 (61) | 74 (59) |  |
| Other, n (%) | 7 (6) | 10 (8) |  |
| Ethnicity |  |  |  |
| Hispanic, n(%) | 5 (4) | 7 (6) | 0.70 |
| Past medical history |  |  |  |
| Diabetes, n (%) | 34 (31) | 46 (37) | 0.32 |
| Liver disease, n (%) | 11 (10) | 25 (20) | 0.03 |
| Renal disease, n (%) | 10 (10) | 24 (20) | 0.03 |
| Physiologic variables |  |  |  |
| Heart rate, beats per minute (IQR) | 100 (84, 113) | 100 (87, 114) | 0.70 |
| Respiratory rate, breaths per minute (IQR) | 20 (16, 24) | 21 (18, 26) | 0.09 |
| Cumulative vasopressor index (IQR) | 4 (3, 8) | 6 (4, 8) | <0.001 |
| Body mass index (IQR) | 28 (25, 36) | 27 (22, 35) | 0.10 |
| Laboratory values |  |  |  |
| White blood count, cells/mm<sup>3</sup> (IQR) | 22.0 (12.3, 28.7) | 16.1 (11.4, 23.7) | 0.24 |
| Platelet count, cells/mm<sup>3</sup> (IQR) | 161 (99, 232) | 129 (65, 210) | 0.02 |
| Creatinine, mg/dL (IQR) | 1.6 (1.1, 2.4) | 2.1 (1.4, 3.0) | 0.003 |
| Total Bilirubin, mg/dL (IQR) | 0.9 (0.5, 1.7) | 1.6 (0.7, 3.7) | <0.001 |
| Clinical lactate, mmol/L (IQR) | 3.1 (2.3, 4.8) | 4.9 (2.7, 8.4) | <0.001 |
| Severity of Illness |  |  |  |
| SOFA score | 10 (8, 12) | 12 (9, 15) | <0.001 |

**Table 2.**
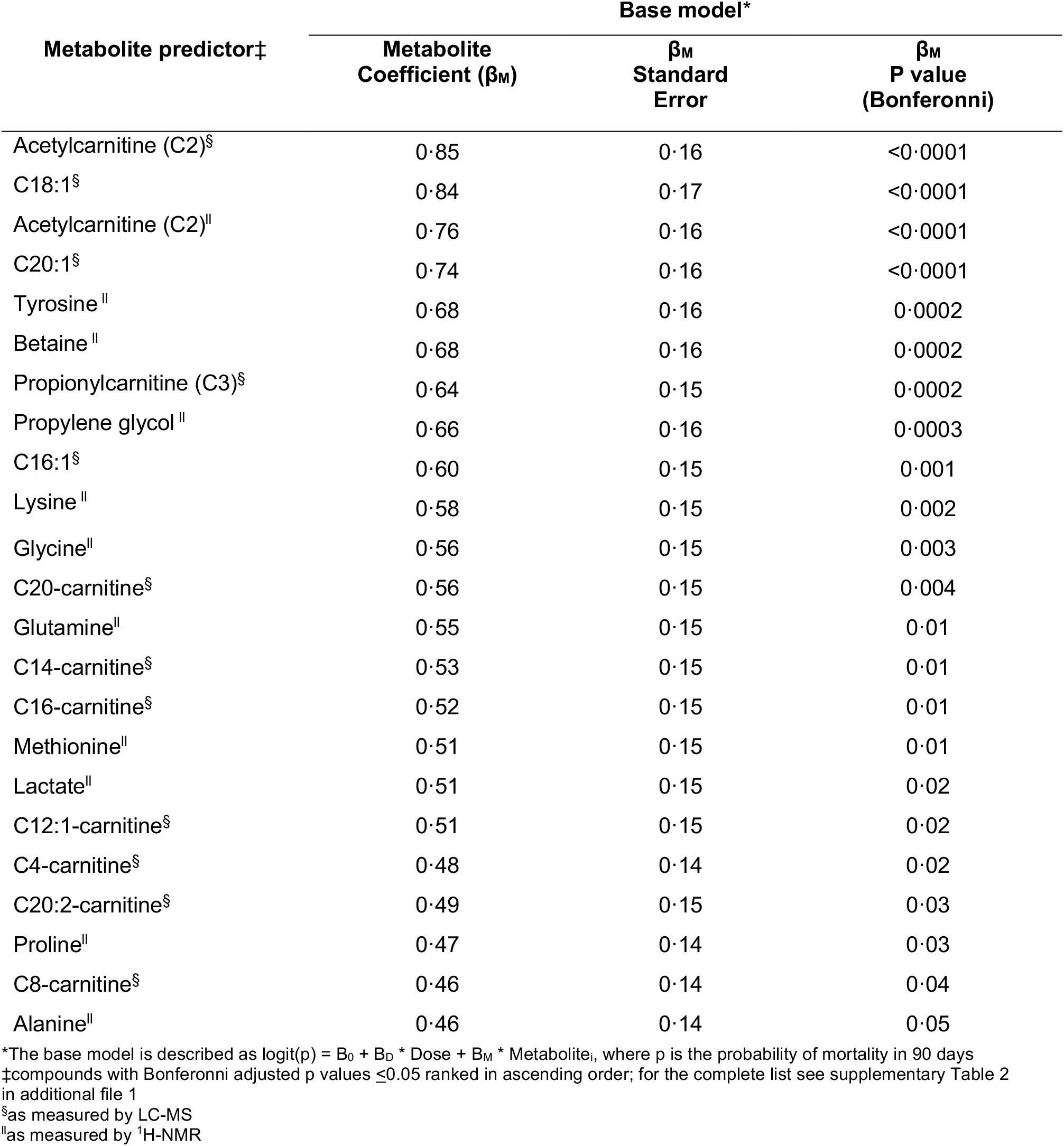
Logistic regression model for the prediction of 90-day mortality adjusted for treatment (L-carnitine dose or placebo)

**Table 3.**
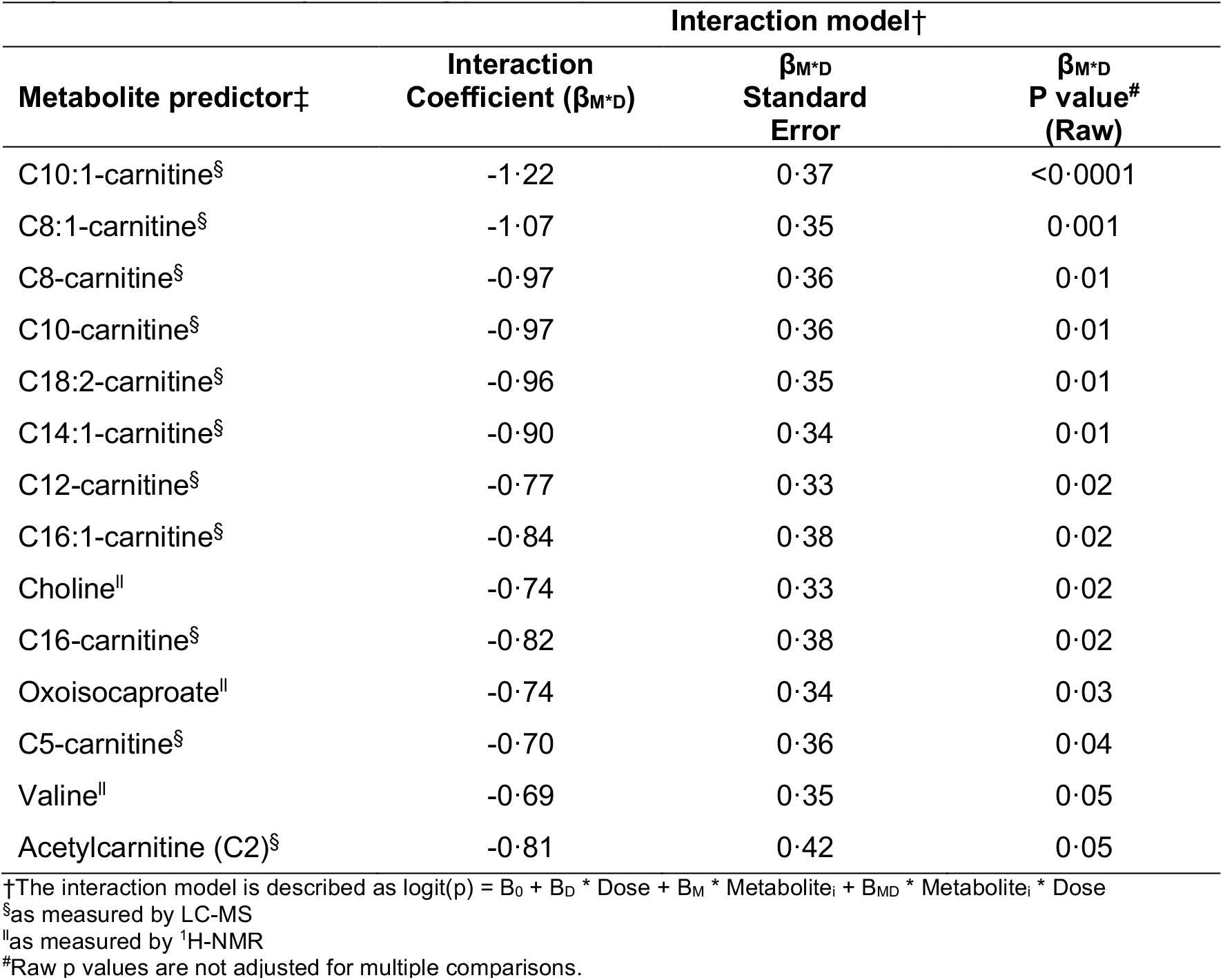
Logistic regression interaction model testing the relationship between metabolite predictors and mortality by treatment (L-carnitine dose or placebo) for the prediction of 90-day mortality ranked by ascending p value up to 0·05

| Metabolite predictor‡ | Interaction model† |  |  |
| --- | --- | --- | --- |
| | Interaction Coefficient ($\beta_{M \cdot D}$) | $\beta_{M \cdot D}$ Standard Error | $\beta_{M \cdot D}$ P value# (Raw) |
| C10:1-carnitine<sup>§</sup> | -1.22 | 0.37 | <0.0001 |
| C8:1-carnitine<sup>§</sup> | -1.07 | 0.35 | 0.001 |
| C8-carnitine<sup>§</sup> | -0.97 | 0.36 | 0.01 |
| C10-carnitine<sup>§</sup> | -0.97 | 0.36 | 0.01 |
| C18:2-carnitine<sup>§</sup> | -0.96 | 0.35 | 0.01 |
| C14:1-carnitine<sup>§</sup> | -0.90 | 0.34 | 0.01 |
| C12-carnitine<sup>§</sup> | -0.77 | 0.33 | 0.02 |
| C16:1-carnitine<sup>§</sup> | -0.84 | 0.38 | 0.02 |
| Choline | -0.74 | 0.33 | 0.02 |
| C16-carnitine<sup>§</sup> | -0.82 | 0.38 | 0.02 |
| Oxoisocaproate | -0.74 | 0.34 | 0.03 |
| C5-carnitine<sup>§</sup> | -0.70 | 0.36 | 0.04 |
| Valine | -0.69 | 0.35 | 0.05 |
| Acetylcarnitine (C2)<sup>§</sup> | -0.81 | 0.42 | 0.05 |

†The interaction model is described as $\text{logit}(p) = B_0 + B_D \cdot \text{Dose} + B_M \cdot \text{Metabolite}_i + B_{MD} \cdot \text{Metabolite}_i \cdot \text{Dose}$
<sup>§</sup>as measured by LC-MS
<sup>||</sup>as measured by <sup>1</sup>H-NMR
#Raw p values are not adjusted for multiple comparisons.

**Table 4.** Significant metabolites^#^ from logistic regression interaction model ranked by descending maximum Z-statistic

| **Metabolite predictor** | **Maximum**<br>**Z-statistic** | **P value** | **FDR (%)<sup>\*</sup>** |
| --- | --- | --- | --- |
| C10:1-carnitine<sup>§</sup> | 3.71 | 0.0002 | 0.30 |
| C8:1-carnitine<sup>§</sup> | 3.50 | 0.0005 | 0.33 |
| C10-carnitine<sup>§</sup> | 3.10 | 0.002 | 0.73 |
| Acetylcarnitine (C2)<sup>§</sup> | 3.06 | 0.002 | 0.73 |
| C8-carnitine<sup>§</sup> | 3.01 | 0.003 | 0.73 |
| C5-carnitine<sup>§</sup> | 2.77 | 0.01 | 1.29 |
| Valine | 2.63 | 0.01 | 1.73 |
| C12-carnitine<sup>§</sup> | 2.58 | 0.01 | 1.73 |
| C14:1-carnitine<sup>§</sup> | 2.54 | 0.01 | 1.75 |
| C18:2-carnitine<sup>§</sup> | 2.50 | 0.01 | 1.75 |
| C16-carnitine<sup>§</sup> | 2.40 | 0.02 | 1.96 |
| C16:1-carnitine<sup>§</sup> | 2.39 | 0.02 | 1.96 |
| Choline | 1.72 | 0.08 | 9.19 |
| Oxoisocaproate | 1.68 | 0.09 | 9.25 |

<sup>#</sup>see Table 3
<sup>§</sup>as measured by LC-MS
<sup>||</sup>as measured by <sup>1</sup>H-NMR
<sup>\*</sup>calculated in MS Excel using the method of Storey, et al<sup>34</sup>

**Figure 2:**
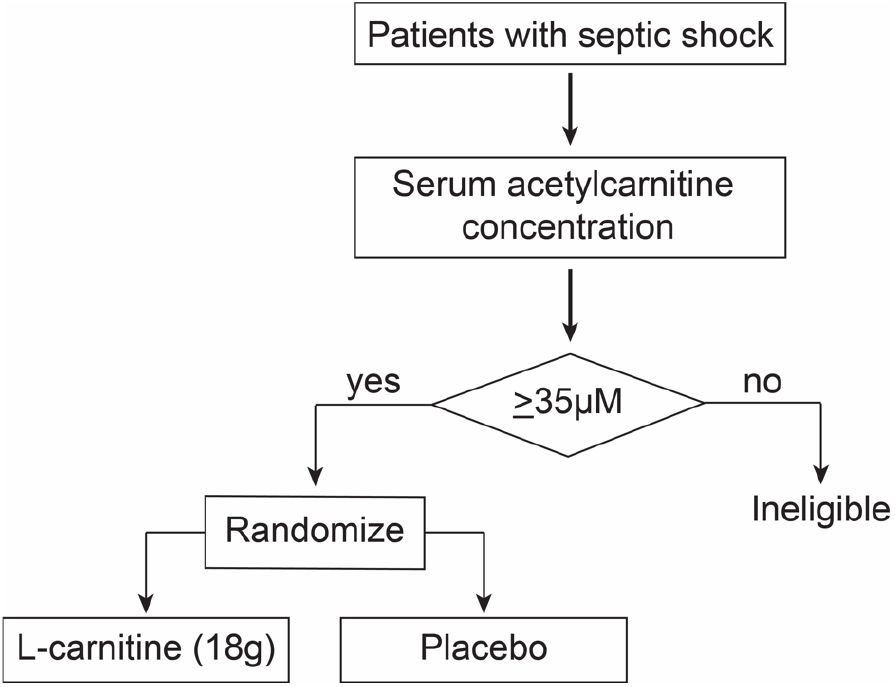
A clinical trial enrichment strategy could optimize clinical trial design for heterogeneous critical illnesses like sepsis. An example of a scheme for a hypothetical phase III clinical trial of supplement L-carnitine for the treatment of septic shock that uses an *a priori* determined acetylcarnitine (C2) threshold concentration to determine whether a patient is enrolled and randomized to receive either L-carnitine (18 g) or placebo.

### Statistical analyses

Descriptive data are reported as means and standard deviations, medians with interquartile ranges, or proportions as appropriate. Differences in categorical outcomes were compared using chi-square tests, while Student t-tests and Wilcoxon rank sum tests were used to compare continuous variables. The aims of our primary analyses were to: 1) determine the relationship between individual metabolites and 90-day mortality; 2) determine if the relationship between a predictive metabolite and mortality depends on treatment allocation; and 3) using metabolites most associated with mortality, determine the optimal (threshold) metabolite level that could be used to identify patients with septic shock most likely to respond favorably to L-carnitine treatment. Collectively, and similar to other secondary analyses or ancillary studies of clinical and observational trials,^21-25^ achievement of these goals would provide clinical proof of concept of a metabolically informed strategy to tackle the heterogeneity of sepsis that also could be used for a predictive enrichment design of a phase III study.^10,11^ Since metabolomics data are on different scales due to varying abundance, in preparation for statistical analyses, data were natural-log transformed and Z-score normalized to have a mean of 0 and a standard deviation of 1.^26,27^ We began our analysis using partial least squares-discriminant analysis (PLS-DA)^28^ to visualize the overall metabolic heterogeneity of the study participants and determine whether there were metabolic differences between sexes and the treatment groups.

We followed PLS-DA by an assessment of the predictive value of individual metabolites on 90-day mortality. To accomplish this, we constructed a series of logistic regression models and adjusted for treatment assignment.^29^ We then tested if the relationship between metabolite predictors and mortality varied across treatment groups using a logistic regression interaction model. Age and SOFA score were considered as covariates in further multivariable modelling.

To test the potential clinical application of our pharmacometabolomics approach, after identifying metabolites strongly related to 90-day mortality that also had a significant interaction with treatment allocation, we aimed to identify the specific concentration or levels of these candidate metabolites that could be used to predict which patients would be most likely to derive a mortality benefit from L-carnitine. To achieve this, we used a grid-search methodology to compute the Z-statistic from the binomial proportion test at every possible threshold metabolite concentration or level.^30^ For this example, since the 18g dose of L-carnitine was the most efficacious in the RACE trial and would be the one most likely to be tested in a phase III trial, we used the Z-statistic to quantify the standardized difference in the proportion of deaths between those patients who received L-carnitine (18 g) and those who received placebo. For this analysis, the metabolite level at each threshold was used as the criterion for inclusion into the proportion test. We then computed a two-sample (binomial) proportion test^31^ which compared the proportion of patients treated with L-carnitine who died by 90 days to those that were treated with placebo. This permitted the identification of metabolite levels associated with a range of Z-statistics including the maximum Z-statistic and their corresponding p values. The Z-statistic simultaneously accounts for the difference in the proportion of patients who died in the treatment versus placebo groups and the sizes of each group, thereby suggesting the most optimal metabolite threshold level. Metabolites were then ranked by descending maximum Z-statistic. Similar approaches have been used by other studies that have sought to identify the responder population in clinical and observational trials.^21-25^ To further illustrate the implications of the use of different metabolite concentrations as predictors of mortality, hazard ratios were calculated using the Mantel-Haenszel method and Kaplan-Meier curves were constructed (log-rank [Mantel-Cox] test). Metabolite concentration cut points were selected according to different trial scenarios and the grid-search analysis described above. All statistical tests except for hazard ratios (Mantel-Haenszel) and log-rank (Mantel-Cox) tests which were done using PRISM, were performed in R studio (R version 3.6.2 (2019-12-12) Copyright 2019 The R Foundation for Statistical Computing) and figures were constructed in R and PRISM (version 8.4.3, June 10, 2020).

## Results

Of the 250 participants randomized in the parent trial, ^1^H-NMR metabolomics and acylcarnitine data were available from 228 and 236 patient serum samples, respectively (see Figure 1 in the supplementary material). We identified and quantified 27 serum metabolites by ^1^H-NMR and 24 acylcarnitines by LC-MS (see Table 1 in the supplementary material).

Representative ^1^H-NMR and LC spectra are shown in Figures 3 and 4 in the supplementary material. All-cause 90-day mortality was 124/236 (52·5%), while 28-day and 1-year mortality were 104/236 (44·1%) and 136/236 (57·6%), respectively. Clinical and demographic variables of the cohort stratified by the primary outcome are summarized in **Table 1**. As expected, patients who died were older and had a higher SOFA score. The PLS-DA plots of the acylcarnitine data and the NMR metabolites by treatment category (supplementary Figure 5A and B) and sex (supplementary Figure 6A and B) illustrate the metabolic heterogeneity of the study cohort and do not demonstrate any significant metabolic distinction between these groups.

We then conducted multivariate logistic regression using L-carnitine dose and metabolites as covariates (base model) and applied a conservative Bonferroni correction for multiple comparisons. The base model identified in 11/27 ^1^H-NMR metabolites and 12/24 acylcarnitines that significantly discriminated 90-day mortality (**Table 2;** the complete list can be found in Table 2 of the supplementary material). We then tested whether the relationship between predictive metabolites and mortality depends on treatment allocation. This was done with the addition of an interaction term between L-carnitine dose and metabolite level (interaction model) which reduced the number of significant metabolites from 23 to 14, of which all but three metabolites were acylcarnitines (**Table 3**; a comprehensive list can be found in Table 3 in the supplementary material); these were not in range to withstand a conservative adjustment (e.g., Bonferroni) for multiple comparisons. In this analysis, a statistically significant and negative interaction term indicates that the predicted probability of 90-day mortality for a given metabolic feature is lower at higher doses of L-carnitine. To determine whether the signals found in the base and interaction models was merely due to factors associated with the risk of death, we controlled for both age^32^ and SOFA score^33^. Several acylcarnitines and choline tolerated this adjustment (see Table 4 in the supplementary material for the full list of metabolites); notably, lactate was not significant in either model (p=0·96 and p=0·22, respectively).

As these findings were not evident in the parent clinical trial and they suggest that there may be a sepsis endotype that may derive a therapeutic benefit from supplement L-carnitine, we hypothesized that a pharmacometabolomics approach may aid in defining this sub-group of patients. To identify the candidate metabolites, we took a hypothesis-generating approach and considered all metabolites with significant (≤ 0·05) unadjusted p values (n=14 in the logistic regression interaction model; **Table 3**) and assessed the Z-statistic of each. Based on this analysis, the metabolites with significant (FDR-corrected p values ≤ 0·05) maximum Z-statistics included a number of acylcarnitines as well as the branched chain amino acid, valine (**Table 4**; also see supplementary Table 5). In addition to the Z-statistic values, to identify candidate metabolites, we also considered the prevalence of the acylcarnitine signal, the known potential of acetylcarnitine (C2) to predict drug responsiveness^15^ and its close metabolic relationship with L-carnitine. Furthermore, the maximum Z-statistic of C12 and C8:1 represented a lower percentage of the clinical cohort than either C5 or acetylcarnitine (C2). As such, we selected acetylcarnitine (C2) as the most viable metabolite candidate to demonstrate the utility of our pharmacometabolomics approach. As examples, we assessed several concentrations of both acetylcarnitine (C2) and valine, including the ones at the maximum Z-statistic, 35 µM (p=0·002; as measured by LC-MS) (**Figure 1**) and 88µM (p=0·009), respectively (also see Figure 7 and Table 5 in the supplementary material). These analyses illustration how a pharmacometabolomics may aid in the design of a precision trial of L-carnitine for the treatment of septic shock using the scheme as illustrated in **Figure 2**.

## Discussion

Our pharmacometabolomics study, CaPS, aimed to identify pre-treatment, sepsis-induced metabolic derangements in survivors and non-survivors treated with L-carnitine. We found that there are likely metabolically distinct groups (endotypes) of patients that do proportionally better when they receive an 18 g dose of supplemental L-carnitine. These findings imply that a precision, clinical trial enrichment strategy using pharmacometabolomics could help combat the heterogeneity of sepsis and drug response, which is known to have contributed to numerous negative clinical studies.^7^

Here we show that a pharmacometabolomics approach identified clinically indistinguishable sepsis endotypes are more likely to derive a mortality benefit from treatment with L-carnitine (18 g), a finding not evident in the metabolically naive parent trial. To accomplish this, we used a metabolomics analysis to capture high abundant polar compounds (quantitative ^1^H-NMR) and acylcarnitines (LC-MS) in serum samples collected from patients enrolled in a phase II clinical trial of L-carnitine therapy.^17^ Using this approach, similar to our prior study^16^, we found a prevalent acylcarnitine signal. From this profile, we selected acetylcarnitine (C2) and valine to illustrate how different threshold concentrations could influence mortality outcome in patients randomized to either placebo of L-carnitine (18 g). Specifically, patients with higher (e.g., ≥ 30µM) acetylcarnitine (C2) levels at enrollment may be more likely to derive a treatment benefit as defined by decreased intermediate term (90-day) mortality; this benefit is maximized at acetylcarnitine (C2) concentrations ≥ 35µM. While severity of illness could contribute to this finding, clinical variables alone do not seem to account for the identification of the drug-responsive endotype since the finding is retained when accounting for factors associated with the risk of death (age and SOFA score, see supplementary Table 4).

Notably, we also found that serum concentrations of the branched chain amino acid, valine, could also be used to identify a mortality benefit of L-carnitine but not to as great an extent as acetylcarnitine (C2). Collectively, these data suggest that there are patients that are in clinically occult subgroups. Should these data be validated, metabolically informed clinical trial design^35^ and ultimately, precision treatment strategies, could represent a new paradigm of sepsis care. These data provide the groundwork and rationale for a pharmacometabolomics directed clinical trial to test L-carnitine therapy efficacy for septic shock using a specific concentration of a key metabolite (e.g., acetylcarnitine (C2)) to guide inclusion criteria (**Figure 2**).

Importantly, the current study also shows that numerous metabolites may have predictive value for sepsis mortality, even after controlling for factors associated with the risk of death (see Table 4 in the supplementary material). These data provide further evidence that sepsis induces broad metabolic disruption that is linked to patient outcomes, corroborating prior studies.^36,37^ Of note, numerous acylcarnitines, including unsaturated acylcarnitines, predicted mortality, suggesting significant disruption in fatty acid metabolic pathways.^37^ Overall, the broad range in disruption of acylcarnitines may reflect differential and variable mobilization of fatty acids,^38^ rather than disruption of a specific enzyme or pathway. We have previously demonstrated this in a smaller cohort of septic shock patients.^16^ Despite this variance, acetylcarnitine (C2) was the most robust predictor of overall sepsis mortality. This corroborates a previous study that identified acetylcarnitine (C2) as being associated with the severity of sepsis-induced organ dysfunction, inflammation and infection.^36^ Acetylcarnitine (C2) also happens to be one of only two compounds (with L-carnitine) detected by both the LC-MS and NMR analytical platforms; regardless of the detection method, it performed similarly in the regression models.

Interestingly, acetylcarnitine (C2) outperformed the more clinically ubiquitous lactate level in predicting sepsis mortality. After correcting for age and SOFA score, lactate was not a significant independent predictor (supplementary Table 4) whereas acetylcarnitine (C2) retained its predictive value following this correction, which suggests the potential for its use as an adjunctive clinical test for risk prognosis. However, as our cohort was highly selected and involved only participants receiving vasopressors (which affect glycolysis and lactate production)^39,40^ who were already resuscitated, it would be inappropriate to interpret these data to imply that lactate does not serve an important role in the early identification and prognosis patients with suspected infection. In particular, serial lactate levels and its clearance rate have been used to assess the adequacy of resuscitation and lactate is included in the sepsis definition.^5,41-43^ Nevertheless, limitations of lactate have been recognized^41^ and, notably, others have demonstrated that acylcarnitines outperform lactate in predicting sepsis mortality.^37^ Our data suggest that acetylcarnitine (C2) may represent a superior risk stratification tool in a selected cohort of fully resuscitated patients undergoing treatment with vasopressor infusions.

We also learned from the CaPS study that pre-treatment serum L-carnitine concentrations did not predict a L-carnitine treatment mortality benefit, suggesting against the hypothesis that serum L-carnitine deficiency drives the response to supplemental L-carnitine in sepsis patients. Rather, in aggregate, these data provide evidence to support the hypothesis that sepsis induces an impairment in the mobilization of acetyl groups. While there may be a number of biologically plausible hypotheses, our findings could be due to sepsis-induced increased intracellular accumulation of acetyl-CoA secondary to its decreased metabolism via the tricarboxylic acid cycle (TCA) or enhanced acetyl-CoA production via fatty acid (beta-oxidation) metabolism (Figure 9 in the supplementary material). Consequently, increases in acetyl-CoA are managed by a number of mechanisms one of which is via the mitochondrial enzyme, carnitine acetyltransferase (EC 2.3.1.7). Carnitine acetyltransferase transfers acetyl groups to carnitine, displacing the hydrogen atom in its hydroxyl group^44^ converting it to the membrane-permeable, acetylcarnitine (C2) (Figure 9 in the supplementary material).

Acetylcarnitine (C2), the shortest of the acylcarnitines, is important because it plays a controlling role over acetyl-CoA on metabolic substrate switching and therefore, enables metabolic flexibility.^44^ As the need for ATP increases, acetyl-CoA is diverted to the TCA cycle. However, in sepsis, the TCA cycle may fail to metabolize these groups resulting in excess acetyl-CoA and subsequent elevation in measured serum acetylcarnitine (C2) concentrations. The elevation in acetylcarnitine (C2) may reflect the ability of L-carnitine to serve as route for the disposal of excess acetyl groups which has been demonstrated in the myocardium^45^ and during exercise.^46^ However, unlike acetylcarnitine (C2), the metabolic link between L-carnitine therapeutic response and BCAA concentrations is less clear. We and others have shown that levels of branched chain amino acids (BCAA) influence sepsis outcome^15,37^ and shock resolution.^47^ It is possible that patients with elevated BCAA blood concentrations represent those with a metabolic reserve that enables them to more efficiently utilize supplemental L-carnitine^48^ but in general, the mechanisms of BCAA signaling and metabolic mechanisms of action are poorly understood.^49^ In aggregate, our findings suggest that the magnitude of sepsis-induced disruptions in energy metabolism may be associated with a therapeutic benefit of L-carnitine. This relationship and the mechanisms that underlie it warrant further interrogation.

Despite the encouraging results of our study, we acknowledge that there are several important weaknesses. We recognize that “real-time” metabolomics is not feasible in clinical practice and that routine measurement of these compounds, including acetylcarnitine (C2), for routine clinical use is not currently available. We also employed a limited, focused metabolomic approach, measuring high abundant polar compounds (^1^H-NMR) and acylcarnitines. We acknowledge that a broad, untargeted approach may have yielded additional compounds predictive of outcomes or treatment response. With our targeted approach, we still made multiple comparisons testing involving over 50 metabolites in this study, which opens the door to false positive findings. Our findings persisted after application of a conservative Bonferroni correction, but we acknowledge that the predictive capacity of acetylcarnitine (C2) and valine, when accounting for interactions between baseline metabolite and treatment assignment (interaction model), was not amenable to correction for multiple comparisons. As such, and given that this was an ancillary study, we acknowledge that any conclusions regarding the accurate prediction of clinical drug responsiveness are only hypothesis generating and will require rigorous prospective testing. We did, however, show how the use of a number of different acetylcarnitine (C2) and valine concentrations would influence the mortality outcome of the RACE trial (**Figure 1** and supplementary Figure 7). These were merely used as examples to illustrate the utility of a pharmacometabolomics approach and despite including almost 250 patients, we acknowledge that our results may overestimate the true effect size and will require validation in an external cohort. Nevertheless, even though these subgroups represent ≤ 50% of the total RACE trial cohort, they highlight the value of a predictive enrichment strategy that could be used to design a phase III clinical trial of L-carnitine supplementation for septic shock.

Importantly, the pharmacometabolomics approach was developed concurrent with the design of the parent trial, and the conceptual model was based on and is consistent with our preliminary work in a unique, though smaller cohort,^15^ strengthening the validity of the findings.

In summary, an ancillary pharmacometabolomics study, CaPS, of the parent clinical trial, RACE, found numerous predictors, independent of intervention, age and SOFA score, for 90-day mortality in septic shock including many acylcarnitines and other metabolites such as tyrosine, betaine, lysine and glycine. We also demonstrate the translational value of the work by showing how the application of a pharmacometabolomics-based clinical trial enrichment strategy, using pre-treatment acetylcarnitine (C2) concentrations as an example, could be used to identify the responder population, a sepsis endotype, that may derive a mortality benefit from L-carnitine supplementation. This represents a unique clinical trial enrichment strategy that could be employed to improve the efficiency of a phase III L-carnitine efficacy study in patients with septic shock^9^ and other emerging therapeutics in heterogeneous critical illnesses. These findings also support the notion that distinct metabolic endotypes contribute to sepsis heterogeneity.

## Data Availability

The metabolomics data sets and R code will be made publicly available at the time of publication of the peer-reviewed manuscript.

https://doi.org/10.21228/M8VX0Z

## Contributors

Authors’ contributions to the study: Study concept and design: MAP, AEJ, KAS; oversight of metabolomics data generation: KAS; LC-MS acylcarnitines data generation: CRE; generation of quantitative NMR metabolomics data: CEM, TLF; analysis data and statistical analyses: MAP, CEG, TSJ, KAS; data interpretation: MAP, CEG, TSJ, AK, KAS; data verification: MAP, TSJ, AEJ, KAS; drafting of the manuscript and its revision for important intellectual content: MAP, CEG, TSJ, CRE, AK, CEM, TLF, AEJ, KAS; final approval of the version submitted for publication: MAP, CEG, TSJ, CRE, AK, CEM, TLF, AEJ, KAS

## Acknowledgements

We acknowledge the contributions of the RACE Trial Investigators group; Nathan I. Shapiro, MD, MPH (Department of Emergency Medicine, Beth Israel Deaconess Medical Center, Boston, MA); Faheem W. Guirgis, MD (Department of Emergency Medicine, University of Florida College of Medicine–Jacksonville, FL); Michael Runyon, MD, MPH (Department of Emergency Medicine, Carolinas Medical Center, Charlotte, NC); Jason Y. Adams, MD (Division of Pulmonary, Critical Care, and Sleep Medicine, Department of Internal Medicine, University of California, Davis, CA); Robert Sherwin, MD (Department of Emergency Medicine, Wayne State University, Detroit, MI); Ryan Arnold, MD (Department of Emergency Medicine, Christiana Care Health System, Wilmington, DE); Brian W. Roberts, MD, MSc (Department of Emergency Medicine, Cooper University Hospital, Cooper Medical School of Rowan University, Camden, NJ); Michael C. Kurz, MD, MS (Department of Emergency Medicine, The University of Alabama School of Medicine at Birmingham, Birmingham, AL); Henry E. Wang, MD, MS (Department of Emergency Medicine, The University of Texas Health Science Center at Houston, Houston, TX); Jeffrey A. Kline, MD (Department of Emergency Medicine, Indiana University School of Medicine, Indianapolis, IN); D. Mark Courtney, MD (Department of Emergency Medicine, Northwestern University, Chicago, IL); Stephen Trzeciak, MD, MPH (Department of Medicine, Cooper University Hospital, Cooper Medical School of Rowan University, Camden, NJ); Sarah A. Sterling, MD (Department of Emergency Medicine, The University of Mississippi Medical Center, Jackson, MS); Utsav Nandi, MD (Department of Emergency Medicine, The University of Mississippi Medical Center, Jackson, MS); Deepti Patki, MS (Department of Emergency Medicine, The University of Mississippi Medical Center, Jackson, MS); Kert Viele, PhD (Berry Consultants, Austin, TX).

