## supplement for "Pharmacometabolomics Identifies Candidate Predictor Metabolites of an L-carnitine Treatment Mortality Benefit in Septic Shock"

On behalf of the RACE Trial Investigators

### Methods

*Patient selection:* Detailed inclusion and exclusion criteria have been previously reported.[1] Briefly, patients presenting with suspected or confirmed infection and meeting consensus criteria for septic shock including administration of continuous vasopressors despite adequate fluid resuscitation, an elevated lactate >2 mmol/L, and a SOFA score of at least 5 were eligible for inclusion. All patients enrolled in the clinical trial were included in the present analysis with the exception of 2 patients who withdrew consent following randomization and 12 that lacked a T0 sample (**supplementary Figure 1**).

*Blood sampling:* Following consent but prior to initiation of therapy, whole blood was drawn into serum separator tubes (SST®; Vacutainer, Becton Dickinson, Franklin Lakes, NJ) and was allowed to clot for at least 30 min at room temperature. A portion of whole blood was aliquoted, while the remainder was centrifuged to yield serum. All samples were then frozen at -80°C within 60 minutes of the initial sample acquisition. Since blood samples were collected from study participants enrolled in a clinical trial, the collection time (T0) of the pre-treatment sample was at the time of enrollment. This was based on when the patient met the study's inclusion criteria and was not anchored to the timing of ICU or hospital admission. This approach has been previously utilized by our group to yield samples appropriate for metabolomics analyses.[2] At the completion of the clinical trial, de-identified serum samples were shipped on dry ice to the University of Michigan (NMR Metabolomics Laboratory) for assay. The

CaPS study was deemed “not regulated” by the University of Michigan’s institutional review board since only de-identified samples and data would be used (HUM00104311).

*Sample handling:* Upon receipt, technical replicate frozen serum samples were transported on dry ice from the NMR Metabolomics Laboratory to the Michigan Regional Comprehensive Metabolomics Research Core. Samples were inventoried, assigned a unique identifier and stored (-80°C) until the time of assay. In preparation for assay, samples were randomized and batched. Samples from a total of 236 of the 250 patients enrolled in the parent clinical trial were assayed; 40 from the dose-response phase of RACE, and 196 from the block randomization phase.[1] Dose response samples were received, processed, and were assayed in advance and separately from the block randomization samples but the randomization schemes were the same for both and the resulting data were combined. For both metabolomics assays, each sample was assigned a random number and samples were ordered by this number for assay. A small volume (50µL) from each sample was reserved for assay of free hemoglobin.[3] The dose response samples were analyzed with Cayman Chemicals hemoglobin colorimetric assay (Ann Arbor, MI; catalog #: 700540), and block randomization samples were analyzed with Arbor Assays hemoglobin high sensitivity detection kit (Ann Arbor, MI; catalog #: K013-HX1). Sample handling was done in accordance with BRISQ guidelines.[4]

*Measurement of Acylcarnitines:* Analysis was performed consistent with our prior methods.[5] Briefly, at the time of analysis, serum samples were thawed on ice and extraction solvent was prepared consisting of a 1:1:1 mixture of methanol:acetonitrile:acetone plus a 1:200 dilution of a stock mixture of stable-isotope labeled acylcarnitine standards dissolved according to manufacturer instructions (NSK-B, Cambridge Isotope Laboratories). Acylcarnitine species were then analyzed without derivatization by RPLC-MS/MS using an Agilent 1200 LC coupled to an Agilent 6410 tandem quadrupole (Santa Clara, CA). A pooled sample generated by combining a small volume of all study samples was injected every 10$^{th}$ run and was used for intra- and inter-batch drift correction as described below. Absolute quantitation by isotope dilution mass spectrometry was performed for acylcarnitine species with exact-matching stable isotope internal standards (L-carnitine, C2, C3, C4, C5, C8, C14, C16) by multiplying the unlabeled / labeled peak area ratio by the known concentration of acylcarnitine in the extraction solvent, then adjusting for dilution of the original serum sample. Other acylcarnitine species were limited to relative quantitation by peak area.

*Quantitative Nuclear Magnetic Resonance (NMR) metabolomics:* Serum samples were prepared by methanol precipitation followed by ultrafiltration as previously described.[6] $^1$H-NMR spectra were acquired at the University of Michigan's Biochemical NMR Core Laboratory on a Varian (now Agilent, Inc., Santa Clara, CA) 11·74 Tesla (500 MHz) NMR spectrometer with a VNMRS console operated by host software VNMRJ 4·0 and equipped with a 5-mm Agilent "One-probe." NMR spectra were recorded using 32 scans of the first increment of a 1 H,1 H-NOESY (commonly referred to as a 1D-NOESY or METNOESY) pulse sequence. Spectra were acquired at a room temperature of 295·45 ± 0·3 K. The NMR pulse sequence was as follows: a 1 s recovery delay, a 990 ms saturation pulse of 80 Hz (gB1) induced field strength empirically centered on the water resonance, 2 calibrated 90° pulses, a mixing time (tmix) of 100 ms, a final 90° pulse, and an acquisition period of 4 s. Optimal excitation pulse widths were obtained by utilizing an array of pulse lengths as previously described.[7] Individuals, blinded to the treatment designation of each sample, analyzed NMR spectra (e.g., carnitine treatment) using Chenomx NMR Suite 8·2 (Edmonton, AB, Canada) software. The Processor module was used to phase shift, baseline correct and excise water from each spectrum. Compounds were identified and relatively quantified using the Profiler module of the software, which allows metabolites to be quantified relative to the area of the internal standard (formate, 50μL of 9·64 mM) added to each sample. Before statistical analysis, data were scaled to correct for differences in initial sample volumes. Only metabolites detected in at least 70% of all samples were considered in the analysis and some samples were excluded because of technical issues (**supplementary Figure 1**).

*Preparation of metabolomics data for statistical analyses*: Patients with any metabolomic data were included in the analyses. Assayed acylcarnitines were detected in all samples.

Because the samples for the acylcarnitine assay were batched and assayed by LC-MS, we assessed and corrected for batch related differences and within-batch intensity using custom-written scripts operating in R. These scripts use local estimate of scatterplot smoothing (LOESS) to correct for within- and between-batch intensity drift in each acylcarnitine species individually, based on peak intensity measured in a pooled QC sample run every 10$^{th}$ sample through all batches.[8,9] For the NMR data set, in preparation for statistical analysis and since concentration values for metabolites that are typically detected can be being missing in the final data set, missing data were replaced with half of the minimum concentration of the respective metabolite across all samples. All detected metabolites and their Kyoto Encyclopedia of Genes and Genomes (KEGG) identification numbers are shown in **supplementary Table 1**. All of the metabolomics data are available at the NIH Common Fund's National Metabolomics Data Repository (NMDR) website, the Metabolomics Workbench, *https://www.metabolomicsworkbench.org* which is supported by NIH grant U2C-DK119886 and where it has been assigned Project ID (accession number ST001319). The data can be accessed directly via it's Project DOIs: (DOI: 10.21228/M8VX0Z).

**Results**

The acquisition of the T0 blood sample occurred at a mean (+SD) of 11(+7) hours from the time of meeting inclusion criteria.

*Serum free hemoglobin*: We assayed all samples for free hemoglobin since we have previously shown that, in serum samples, it may contribute to a distortion in the concentration of certain metabolites particularly if the hemoglobin concentration is greater than 0·1 g/dL.[3] All but one (0·2g/dL) of the assayed samples had a free hemoglobin concentration below 0·1 g/dL (data not shown).

| **Supplementary Table 1**: Acylcarnitines and $^{1}$H-NMR Detected and Quantified Metabolites | | | |
|---|---|---|---|
| **Acylcarnitines** | | **$^{1}$H-NMR Metabolites$^{\#}$** | |
| Compound Name | KEGG ID$^{+}$ | Compound Name | KEGG ID$^{+}$ |
| **L-Carnitine** | C00318 | 2-Hydroxybutyrate | C05984 |
| **C2-carnitine** | C02571 | 2-Oxoisocaproate | C00233 |
| **C3-carnitine** | C03017 | 3-Hydroxybutyrate | C01089 |
| **C4-carnitine** | C02862 | Acetylcarnitine (C2) | C02571 |
| **C5-carnitine** | HMDB0013128* | Alanine | C00041 |
| C6-carnitine | HMDB0000756* | Betaine | C00719 |
| C8:1-carnitine | HMDB0013324* | Carnitine | C00318 |
| **C8-carnitine** | C02838 | Choline | C00114 |
| C10:1-carnitine | HMDB0240585* | Citrate | C00158 |
| C10-carnitine | C03299 | Creatine | C00300 |
| C12:1-carnitine | HMDB0013326* | Creatinine | C00791 |
| C12-carnitine | HMDB0002250* | Glucose | C00221 |
| C14:1-carnitine | HMDB0002014* | Glutamine | C00064 |
| **C14-carnitine** | HMDB0005066* | Glycine | C00037 |
| C16:1-carnitine | HMDB0006317* | Histidine | C00135 |
| C20:4-carnitine | HMDB0006455* | Isoleucine | C00407 |
| C18:2-carnitine | HMDB0006469* | Lactate | C00186 |
| C20:3-carnitine | N/A | Leucine | C00123 |
| **C16-carnitine** | C02990 | Lysine | C00047 |
| C18:1-carnitine | HMDB0006464* | Methionine | C00073 |
| C20:2-carnitine | N/A | Ornithine | C00077 |
| C18-carnitine | HMDB0000848* | Phenylalanine | C00079 |
| C20:1-carnitine | N/A | Proline | C00148 |
| C20-carnitine | HMDB0006455* | Propylene glycol | C00583 |
| | | Pyruvate | C00022 |
| | | Tyrosine | C00082 |
| | | Valine | C00183 |

+Kyoto Encyclopedia of Genes and Genomes (KEGG) identification number
*Human metabolomics database (HMDB) identification number; KEGG ID not available
N/A = neither a KEGG nor HMDB ID is available
**Bolded** acylcarnitines were quantified using an exact-matching stable isotope internal standard; all others were relatively quantified based on peak-area
$^{\#}$NMR metabolites were quantified from the addition of a known concentration of an internal standard (formate) that was added to each sample

**Supplementary Table 2**: Logistic regression base model for the prediction of 90-day mortality (all metabolites) ranked by ascending Bonferroni-corrected p value

| | Base model* | | |
|---|---|---|---|
| **Metabolite predictor** | **Metabolite Coefficient ($\beta_M$)** | **$\beta_M$ Standard Error** | **$\beta_M$ P value (Bonferroni)** |
| Acetylcarnitine (C2)[§] | 0·85 | 0·16 | <0·0001 |
| C18:1-carnitine[§] | 0·84 | 0·17 | <0·0001 |
| Acetylcarnitine (C2)[‖] | 0·76 | 0·16 | <0·0001 |
| C20:1-carnitine[§] | 0·74 | 0·16 | <0·0001 |
| Tyrosine [‖] | 0·68 | 0·16 | 0·0002 |
| Betaine [‖] | 0·68 | 0·16 | 0·0002 |
| Propionylcarnitine (C3)[§] | 0·64 | 0·15 | 0·0002 |
| Propylene glycol[‖] | 0·66 | 0·16 | 0·0003 |
| C16:1-carnitine[§] | 0·60 | 0·15 | 0·001 |
| Lysine[‖] | 0·58 | 0·15 | 0·002 |
| Glycine[‖] | 0·56 | 0·15 | 0·003 |
| C20-carnitine[§] | 0·56 | 0·15 | 0·004 |
| Glutamine[‖] | 0·55 | 0·15 | 0·006 |
| C14-carnitine[§] | 0·53 | 0·15 | 0·007 |
| C16-carnitine[§] | 0·52 | 0·15 | 0·01 |
| Methionine[‖] | 0·51 | 0·15 | 0·01 |
| Lactate[‖] | 0·51 | 0·15 | 0·02 |
| C12:1-carnitine[§] | 0·51 | 0·15 | 0·02 |
| C4-carnitine[§] | 0·48 | 0·14 | 0·02 |
| C20:2-carnitine[§] | 0·49 | 0·15 | 0·03 |
| Proline[‖] | 0·47 | 0·14 | 0·03 |
| C8-carnitine[§] | 0·46 | 0·14 | 0·04 |
| Alanine[‖] | 0·46 | 0·14 | 0·05 |
| C18-carnitine[§] | 0·44 | 0·15 | 0·09 |
| Creatinine[‖] | 0·43 | 0·14 | 0·09 |
| C10-carnitine[§] | 0·41 | 0·14 | 0·13 |
| C5-carnitine[§] | 0·41 | 0·14 | 0·14 |
| Phenylalanine[‖] | 0·39 | 0·15 | 0·29 |
| Citrate[‖] | 0·36 | 0·14 | 0·45 |
| Carnitine[‖] | 0·35 | 0·14 | 0·52 |
| Histidine[‖] | 0·35 | 0·14 | 0·52 |
| C10:1-carnitine[§] | 0·34 | 0·14 | 0·58 |
| C6-carnitine[§] | 0·34 | 0·14 | 0·59 |
| C12-carnitine[§] | 0·31 | 0·14 | 1·00 |

| | | | |
|---|---|---|---|
| C18:2-carnitine§ | 0·30 | 0·14 | 1·00 |
| Ornithine‖ | 0·29 | 0·14 | 1·00 |
| C14:1-carnitine§ | 0·29 | 0·14 | 1·00 |
| Creatine‖ | 0·25 | 0·14 | 1·00 |
| Carnitine§ | 0·25 | 0·14 | 1·00 |
| Pyruvate‖ | 0·23 | 0·14 | 1·00 |
| Leucine‖ | 0·21 | 0·14 | 1·00 |
| Choline‖ | 0·20 | 0·14 | 1·00 |
| Valine‖ | 0·18 | 0·14 | 1·00 |
| 2-hydroxybutyrate‖ | 0·18 | 0·14 | 1·00 |
| C20:3-carnitine§ | 0·17 | 0·14 | 1·00 |
| C8:1-carnitine§ | 0·16 | 0·13 | 1·00 |
| C20:4-carnitine§ | 0·11 | 0·14 | 1·00 |
| 3-hydroxybutyrate‖ | 0·10 | 0·14 | 1·00 |
| Isoleucine‖ | 0·08 | 0·13 | 1·00 |
| Glucose‖ | 0·07 | 0·14 | 1·00 |
| Oxoisocaproate‖ | -0·06 | 0·14 | 1·00 |

*The base model is described as logit(p) = $B_0 + B_D$ * Dose + $B_M$ * $Metabolite_i$, where p is the probability of mortality in 90 days

§as measured by LC-MS

‖as measured by $^1$H-NMR

**Supplementary Table 3**: Logistic regression interaction model for the prediction of 90-day mortality (all metabolites) ranked by ascending p value

| | Interaction model[†] | | |
|---|---|---|---|
| **Metabolite predictor** | **Interaction Coefficient ($\beta_{M*D}$)** | **$\beta_{M*D}$ Standard Error** | **$\beta_{M*D}$ P value[‡] (Raw)** |
| C10:1-carnitine[§] | -1·22 | 0·37 | <0·0001 |
| C8:1-carnitine[§] | -1·07 | 0·35 | 0·001 |
| C8-carnitine[§] | -0·97 | 0·36 | 0·01 |
| C10-carnitine[§] | -0·97 | 0·36 | 0·01 |
| C18:2-carnitine[§] | -0·96 | 0·35 | 0·01 |
| C14:1-carnitine[§] | -0·90 | 0·34 | 0·01 |
| C12-carnitine[§] | -0·77 | 0·33 | 0·02 |
| C16:1-carnitine[§] | -0·84 | 0·38 | 0·02 |
| Choline[‖] | -0·74 | 0·33 | 0·02 |
| C16-carnitine[§] | -0·82 | 0·38 | 0·02 |
| Oxoisocaproate[‖] | -0·74 | 0·34 | 0·03 |
| C5-carnitine[§] | -0·70 | 0·36 | 0·04 |
| Valine[‖] | -0·69 | 0·35 | 0·05 |
| Acetylcarnitine (C2)[§] | -0·81 | 0·42 | 0·05 |
| C12:1-carnitine[§] | -0·70 | 0·37 | 0·06 |
| Isoleucine[‖] | -0·57 | 0·31 | 0·06 |
| Carnitine[‖] | -0·64 | 0·36 | 0·07 |
| Glycine[‖] | -0·62 | 0·35 | 0·08 |
| C14-carnitine[§] | -0·56 | 0·35 | 0·10 |
| C18-carnitine[§] | -0·56 | 0·36 | 0·11 |
| Proline[‖] | -0·52 | 0·34 | 0·12 |
| Acetylcarnitine (C2)[‖] | -0·57 | 0·40 | 0·14 |
| C6-carnitine[§] | -0·47 | 0·33 | 0·14 |
| Lactate[‖] | 0·52 | 0·37 | 0·15 |
| Propylene glycol[‖] | -0·54 | 0·39 | 0·16 |
| Citrate[‖] | -0·47 | 0·34 | 0·16 |
| Leucine[‖] | -0·45 | 0·34 | 0·17 |
| Carnitine[§] | -0·44 | 0·33 | 0·18 |
| C20:4-carnitine[§] | 0·44 | 0·44 | 0·21 |
| C20:3-carnitine[§] | -0·39 | 0·32 | 0·22 |
| Methionine[‖] | -0·40 | 0·34 | 0·23 |
| Propionylcarnitine (C3)[§] | -0·42 | 0·36 | 0·24 |
| Pyruvate[‖] | 0·36 | 0·33 | 0·28 |

| | | | |
|---|---|---|---|
| C20:2-carnitine§ | -0·37 | 0·36 | 0·30 |
| 2-hydroxybutyrate‖ | -0·30 | 0·31 | 0·34 |
| 3-hydroxybutyrate‖ | -0·30 | 0·32 | 0·34 |
| C20-carnitine§ | -0·27 | 0·38 | 0·47 |
| Phenylalanine‖ | -0·23 | 0·35 | 0·51 |
| Creatine‖ | -0·18 | 0·33 | 0·58 |
| Tyrosine ‖ | 0·16 | 0·39 | 0·68 |
| Ornithine‖ | 0·13 | 0·32 | 0·69 |
| Alanine‖ | -0·13 | 0·35 | 0·71 |
| Creatinine‖ | -0·10 | 0·33 | 0·75 |
| C20:1-carnitine§ | -0·10 | 0·39 | 0·80 |
| C18:1-carnitine§ | -0·10 | 0·39 | 0·80 |
| Glucose‖ | 0·06 | 0·33 | 0·86 |
| C4-carnitine§ | -0·03 | 0·34 | 0·92 |
| Histidine‖ | 0·03 | 0·34 | 0·93 |
| Glutamine‖ | -0·03 | 0·37 | 0·94 |
| Betaine ‖ | 0·02 | 0·38 | 0·96 |
| Lysine‖ | 0·01 | 0·35 | 0·97 |

†The interaction model is described as $\text{logit}(p) = B_0 + B_D * \text{Dose} + B_M * \text{Metabolite}_i + B_{MD} * \text{Metabolite}_i * \text{Dose}$
§as measured by LC-MS
‖as measured by $^1$H-NMR
#Raw p values are not adjusted for multiple comparisons.

**Supplementary Table 4**: Logistic regression models for the prediction of 90-day mortality adjusted for age and Sequential Organ Failure Assessment (SOFA) Score

| | Base model* | | | Interaction model† | | |
|---|---|---|---|---|---|---|
| **Metabolite predictor** | **Metabolite Coefficient ($\beta_M$)** | **$\beta_M$ Standard Error** | **$\beta_M$ P value‡ (Bonferonni)** | **Interaction Coefficient ($\beta_{M*D}$)** | **$\beta_{M*D}$ Standard Error** | **$\beta_{M*D}$ P value‡ (Raw)** |
| C18:1-carnitine§ | 0·73 | 0·18 | 0·0004 | -0·02 | 0·41 | 0·96 |
| Acetylcarnitine (C2) § | 0·70 | 0·17 | 0·001 | -0.82 | 0·43 | 0·05 |
| C20:1-carnitine § | 0·67 | 0·18 | 0·002 | -0·05 | 0·41 | 0·90 |
| Betaine‖ | 0·62 | 0·18 | 0·009 | -0·12 | 0·41 | 0·76 |
| Acetylcarnitine (C2)§ | 0·57 | 0·17 | 0·02 | -0·46 | 0·41 | 0·25 |
| Tyrosine ‖ | 0·55 | 0·17 | 0·02 | 0·10 | 0·40 | 0·79 |
| C16-carnitine§ | 0·53 | 0·16 | 0·03 | -0·85 | 0·41 | 0·03 |
| C16:1-carnitine§ | 0·53 | 0·16 | 0·03 | -0·90 | 0·41 | 0·02 |
| C20-carnitine§ | 0·53 | 0·16 | 0·03 | -0·17 | 0·40 | 0·67 |
| Propionylcarnitine (C3)§ | 0·49 | 0·16 | 0·06 | -0·35 | 0·38 | 0·35 |
| C18-carnitine§ | 0·46 | 0·16 | 0·11 | -0·49 | 0·39 | 0·20 |
| Lysine‖ | 0·47 | 0·16 | 0·12 | 0·00 | 0·37 | 0·99 |
| C20:2-carnitine§ | 0·45 | 0·16 | 0·16 | -0·39 | 0·38 | 0·29 |
| Glycine‖ | 0·47 | 0·16 | 0·16 | -0·69 | 0·38 | 0·07 |
| Glutamine‖ | 0·47 | 0·17 | 0·17 | -0·12 | 0·40 | 0·76 |
| C14-carnitine§ | 0·44 | 0·15 | 0·18 | -0·57 | 0·37 | 0·12 |
| C12:1-carnitine§ | 0·44 | 0·16 | 0·25 | -0·83 | 0·40 | 0·03 |
| Proline‖ | 0·41 | 0·16 | 0·36 | -0·57 | 0·37 | 0·11 |
| Propylene glycol‖ | 0·44 | 0·17 | 0·36 | -0·39 | 0·40 | 0·33 |
| Methionine‖ | 0·40 | 0·16 | 0·46 | -0·39 | 0·36 | 0·26 |
| Creatinine‖ | 0·35 | 0·15 | 0·92 | -0·03 | 0·35 | 0·94 |
| Lactate‖ | 0·37 | 0·16 | 0·96 | 0·47 | 0·39 | 0·22 |
| C10--carnitine§ | 0·34 | 0·15 | 0·98 | -0·94 | 0·37 | 0·01 |
| Carnitine§ | 0·08 | 0·15 | 1·00 | -0·35 | 0·36 | 0·32 |
| C4-carnitine§ | 0·31 | 0·15 | 1·00 | 0·16 | 0·36 | 0·66 |
| C5-carnitine§ | 0·24 | 0·15 | 1·00 | -0·57 | 0·38 | 0·12 |
| C6-carnitine § | 0·18 | 0·15 | 1·00 | -0·37 | 0·34 | 0·28 |
| C8:1-carnitine§ | 0·09 | 0·14 | 1·00 | -1·06 | 0·37 | 0·003 |
| C8--carnitine§ | 0·31 | 0·15 | 1·00 | -0·87 | 0·37 | 0·02 |
| C10:1-carnitine§ | 0·26 | 0·15 | 1·00 | -1·23 | 0·39 | 0·001 |
| C12-carnitine§ | 0·30 | 0·15 | 1·00 | -0·88 | 0·37 | 0·01 |
| C14:1-carnitine§ | 0·27 | 0·15 | 1·00 | -0·98 | 0·37 | 0·01 |

| | | | | | | |
|---|---|---|---|---|---|---|
| C20:4-carnitine§ | -0·06 | 0·16 | 1·00 | 0·86 | 0·78 | 0·05 |
| C18:2-carnitine§ | 0·28 | 0·15 | 1·00 | -0·93 | 0·38 | 0·01 |
| C20:3-carnitine§ | 0·20 | 0·15 | 1·00 | -0·34 | 0·34 | 0·33 |
| 2-Hydroxybutyrate‖ | 0·11 | 0·15 | 1·00 | -0·19 | 0·33 | 0·56 |
| Oxoisocaproate‖ | 0·00 | 0·15 | 1·00 | -0·42 | 0·38 | 0·27 |
| 3-Hydoxybutyrate‖ | 0·11 | 0·15 | 1·00 | -0·29 | 0·34 | 0·40 |
| Alanine‖ | 0·35 | 0·16 | 1·00 | -0·15 | 0·37 | 0·69 |
| Carnitine‖ | 0·20 | 0·15 | 1·00 | -0·61 | 0·38 | 0·10 |
| Choline‖ | 0·12 | 0·15 | 1·00 | -0·68 | 0·35 | 0·05 |
| Citrate‖ | 0·28 | 0·16 | 1·00 | -0·47 | 0·37 | 0·21 |
| Creatine‖ | 0·10 | 0·15 | 1·00 | -0·24 | 0·36 | 0·50 |
| Glucose‖ | 0·04 | 0·15 | 1·00 | 0·04 | 0·35 | 0·91 |
| Histidine‖ | 0·24 | 0·15 | 1·00 | -0·14 | 0·36 | 0·70 |
| Isoleucine‖ | 0·09 | 0·15 | 1·00 | -0·45 | 0·33 | 0·17 |
| Leucine‖ | 0·23 | 0·15 | 1·00 | -0·36 | 0·36 | 0·32 |
| Ornithine‖ | 0·29 | 0·15 | 1·00 | 0·24 | 0·34 | 0·49 |
| Phenylalanine‖ | 0·34 | 0·16 | 1·00 | -0·10 | 0·38 | 0·80 |
| Pyruvate‖ | 0·13 | 0·15 | 1·00 | 0·41 | 0·35 | 0·24 |
| Valine‖ | 0·21 | 0·15 | 1·00 | -0·52 | 0·38 | 0·17 |

*The base model is described as logit(p) = $B_0 + B_D * Dose + B_M * Metabolite_i + B_{SOFA} * SOFA + B_{Age} * Age$ where p is the probability of mortality in 90 days

†The interaction model is described as logit(p) = $B_0 + B_D * Dose + B_M * Metabolite_i + B_{MD} * Metabolite_i * Dose + B_{SOFA} * SOFA + B_{Age} * Age$

‡ p-values were calculated upon comparison to the appropriate nested model using the likelihood ratio test as described in the supplement methods section; raw p values are not adjusted for multiple comparisons.

§as measured by LC-MS

‖as measured by $^1$H-NMR

**Supplementary Table 5**: Threshold concentrations of candidate metabolites from the interaction regression model that also have a significant (FDR ≤ 0.05%) maximum Z-statistic.

| | | | |
|---|---|---|---|
| ***C10:1 carnitine*** | | | |
| **Threshold** (concentration, μM) | **Recall** (Percent of 18g or placebo-treated patients) | Z-Statistic† | P value |
| 0·39 | ~50 | 2·39 | 0·02 |
| 0·44 | ~40 | 3·126 | 0·001 |
| 0·47 | ~30 | 3·71[#] | 0·0002 |
| 0·55 | ~20 | 2·33 | 0·02 |
| ***C8:1 carnitine*** | | | |
| **Threshold** (concentration, μM) | **Recall** (Percent of 18g or placebo-treated patients) | Z-Statistic† | P value |
| 0·46 | ~50 | 2·01 | 0·04 |
| 0·56 | ~40 | 2·23 | 0·03 |
| 0·69 | ~30 | 2·97 | 0·003 |
| 0·81 | ~20 | 3·44 | 0·0006 |
| 0·84 | ~16 | 3·50[#] | 0·0005 |
| ***C10 carnitine*** | | | |
| **Threshold** (concentration, μM) | **Recall** (Percent of 18g or placebo-treated patients) | Z-Statistic† | P value |
| 0·42 | ~50 | 2·13 | 0·03 |
| 0·51 | ~40 | 2·46 | 0·01 |
| 0·58 | ~30 | 2·93 | 0·003 |
| 0·66 | ~25 | 3·10[#] | 0·002 |
| 0·73 | ~20 | 2·88 | 0·004 |
| ***Acetylcarnitine (C2)**** [+] | | | |
| **Threshold** (concentration, μM) | **Recall** (Percent of 18g or placebo-treated patients) | Z-Statistic† | P value |
| 18 | ~50 | 0·152 | 0·88 |
| 21 | ~40 | 0·990 | 0·32 |
| 28 | ~30 | 2·33 | 0·02 |
| 35 | ~20 | 3·06[#] | 0·002 |
| ***C8 carnitine*** | | | |
| **Threshold** (concentration, μM) | **Recall** (Percent of 18g or placebo-treated patients) | Z-Statistic† | P value |
| 0·23 | ~50 | 2·03 | 0·04 |
| 0·28 | ~40 | 2·47 | 0·01 |
| 0·32 | ~30 | 3·01[#] | 0·003 |
| 0·45 | ~20 | 2·85 | 0·004 |
| ***C5 carnitine*** | | | |
| **Threshold** (concentration, μM) | **Recall** (Percent of 18g or placebo-treated patients) | Z-Statistic† | P value |
| 0·19 | ~50 | 1·05 | 0·29 |
| 0·22 | ~40 | 1·75 | 0·08 |
| 0·26 | ~30 | 2·03 | 0·04 |
| 0·27 | ~26 | 2·77[#] | 0·006 |
| 0·29 | ~20 | 2·50 | 0·01 |

| ***Valine**** | | | |
|---|---|---|---|
| **Threshold** (concentration, μM) | **Recall** (Percent of 18g or placebo-treated patients) | Z-Statistic† | P value |
| 88 | ~50 | 2·62[#] | 0·009 |
| 97 | ~40 | 2·34 | 0·02 |
| 113 | ~30 | 2·18 | 0·03 |
| 130 | ~20 | 2·11 | 0·03 |
| ***C12 Carnitine*** | | | |
| **Threshold** (concentration, μM) | **Recall** (Percent of 18g or placebo-treated patients) | Z-Statistic† | P value |
| 0·040 | ~50 | 1·41 | 0·16 |
| 0·050 | ~40 | 1·59 | 0·11 |
| 0·068 | ~30 | 1·66 | 0·10 |
| 0·094 | ~20 | 1·95 | 0·05 |
| 0·102 | ~13 | 2·58[#] | 0·01 |
| ***C14:1 carnitine*** | | | |
| **Threshold** (concentration, μM) | **Recall** (Percent of 18g or placebo-treated patients) | Z-Statistic† | P value |
| 0·045 | ~50 | 2·13 | 0·03 |
| 0·057 | ~40 | 2·19 | 0·03 |
| 0·079 | ~30 | 1·74 | 0·08 |
| 0·110 | ~20 | 2·32 | 0·02 |
| 0·262 | ~6 | 2·54[#] | 0·01 |
| ***C18:2 carnitine*** | | | |
| **Threshold** (concentration, μM) | **Recall** (Percent of 18g or placebo-treated patients) | Z-Statistic† | P value |
| 0·060 | ~50 | 2·00 | 0·05 |
| 0·072 | ~40 | 2·17 | 0·03 |
| 0·089 | ~30 | 1·66 | 0·01 |
| 0·107 | ~20 | 1·66 | 0·01 |
| 0·163 | ~9 | 2·50[#] | 0·01 |
| ***C16 carnitine*** | | | |
| **Threshold** (concentration, μM) | **Recall** (Percent of 18g or placebo-treated patients) | Z-Statistic† | P value |
| 0·043 | ~54 | 2·40[#] | 0·02 |
| 0·055 | ~40 | 1·70 | 0·09 |
| 0·070 | ~30 | 1·70 | 0·09 |
| 0·100 | ~20 | 1·682 | 0·07 |
| ***C16:1 Carnitine*** | | | |
| **Threshold** (concentration, μM) | **Recall** (Percent of 18g or placebo-treated patients) | Z-Statistic† | P value |
| 0·029 | ~50 | 2·39[#] | 0·02 |
| 0·036 | ~40 | 1·91 | 0·06 |
| 0·045 | ~30 | 1·76 | 0·08 |
| 0·064 | ~20 | 2·21 | 0·03 |

†with the exception of the maximum Z-statistic, the Z-statistic represents the maximum value within (±) a 2% range of the associated recall
*metabolite concentrations were rounded to the nearest whole number
[+]as measured by liquid chromatography-mass spectrometry
[^]as measured by nuclear magnetic resonance spectroscopy
[#]maximum Z-statistic

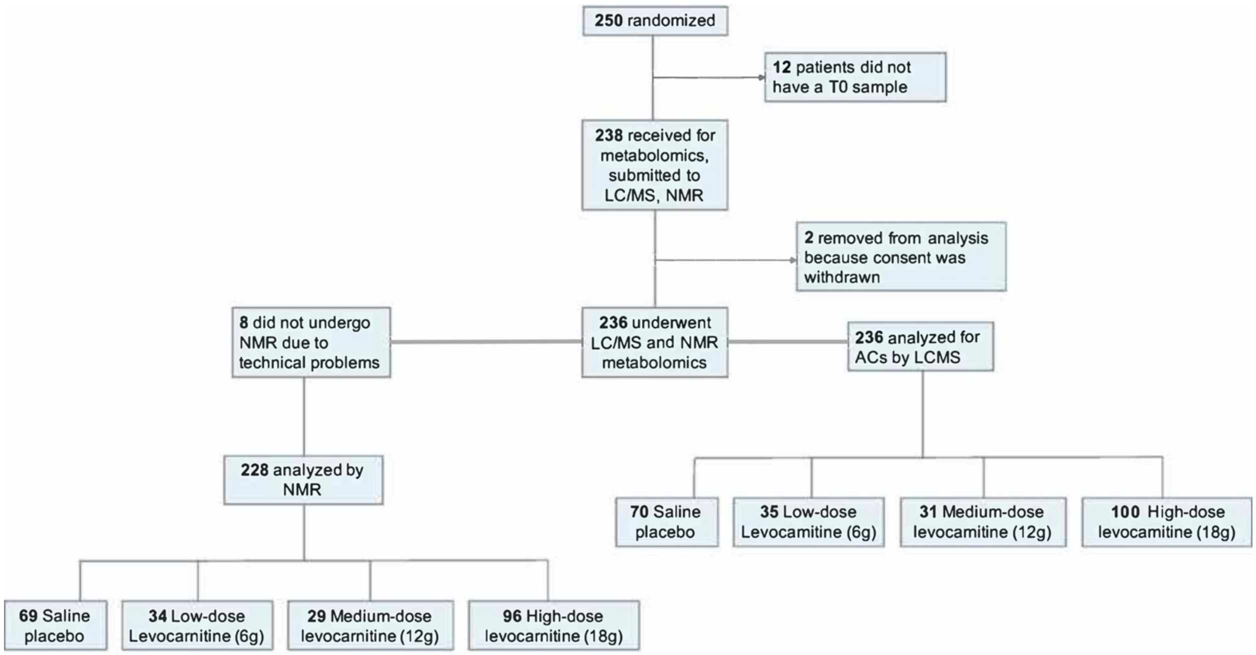


**Supplementary Figure 1**: Consort diagram of patient samples for metabolomics analysis· A total of 250 patients were enrolled and randomized in the Rapid Administration of Carnitine in Sepsis (RACE) trial [1]. Twelve participants did not have a T0 blood sample because the sample was not collected. Of the total of 238 samples that were received for metabolomics analysis, two were excluded from analysis because of withdrawal of consent. Of the 236 samples that were submitted for NMR analysis, there were 8 that had insufficient volume or did not extract properly. The final data set are represented by 228 samples with NMR metabolomics data and 236 with acylcarnitine data.

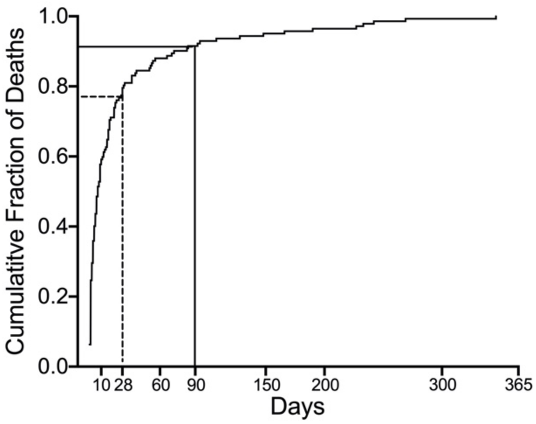


**Supplementary Figure 2**: The cumulative fraction of deaths of the patients who died in the Rapid Administration of Carnitine in Sepsis (RACE) clinical trial. The dotted line corresponds to the cumulative fraction of deaths at 28 days (0·78) and the solid line corresponds to the cumulative fraction of deaths at 90 days (0·91).

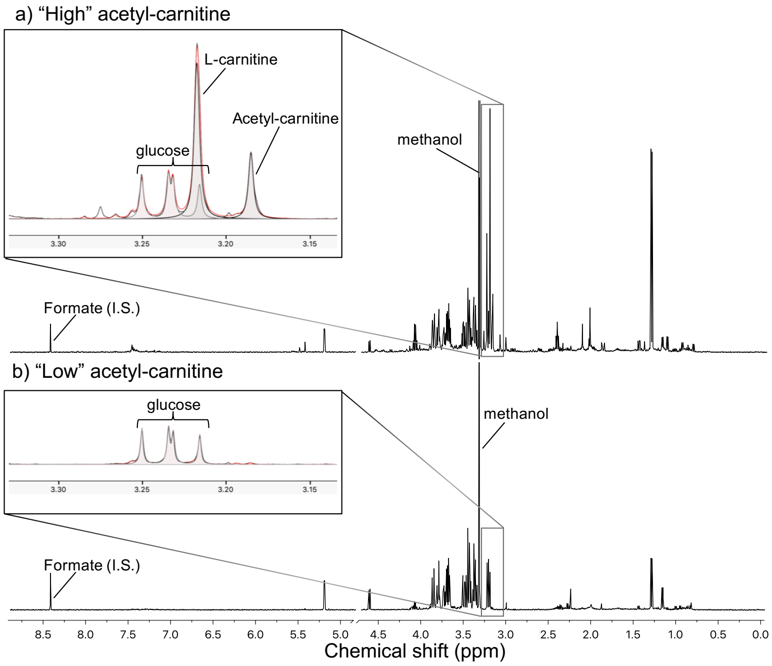


**Supplementary Figure 3**: Representative proton ([1]H) nuclear magnetic resonance (NMR) spectra of (A) a serum sample from a patient with “high” acetylcarnitine (C2) concentration ($\geq$ 35 µM) and (B) a “low” acetylcarnitine (C2) concentration (< 35µM). Formate was added to each sample as the internal standard (I.S.). The compounds (glucose, L-carnitine and acetylcarnitine) are the most abundant in the spectral region shown in the insets. Methanol is introduced into samples via the precipitation process.

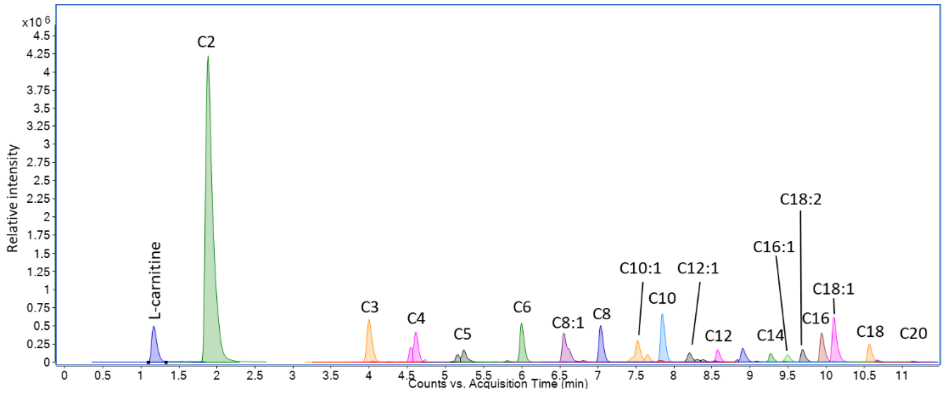


**Supplementary Figure 4**: A representative liquid chromatography-mass spectroscopy chromatogram of acylcarnitines in a pooled plasma sample. The chromatogram was generated by overlaying multiple reaction monitoring transitions for all detected acylcarnitine species; major species were identified by peak labels in the chromatogram. Isotopically-labeled internal standard peaks are omitted from this chromatogram for clarity, but essentially co-eluted with the matching unlabeled acylcarnitine species.

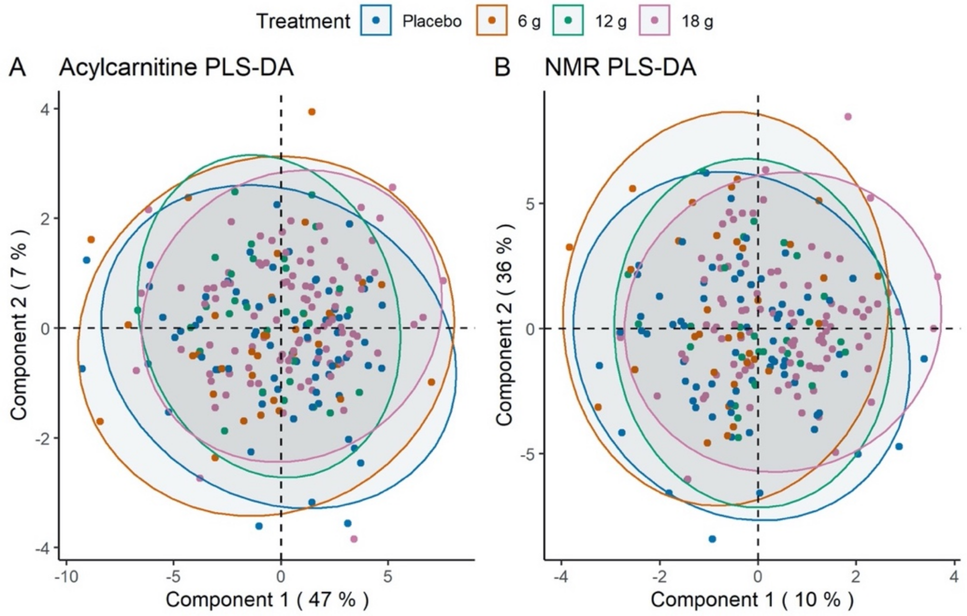


**Supplementary Figure 5**: **Partial Least Squares-Discriminant Analysis (PLS-DA) does not discriminate acylcarnitines and nuclear magnetic resonance (NMR)-detected metabolites in patients with septic shock treated with L-carnitine (6, 12, or 18 g) or placebo (saline).** PLS-DA (and associated multivariate t distribution) of serum acylcarnitines (**A**) and NMR-detected metabolites (**B**) from the 236 and 228 patients, respectively, of the 250 patients enrolled in the Rapid Administration of Carnitine (RACE) in Sepsis clinical trial.[1] The results highlight the metabolic heterogeneity of the cohort and demonstrate that there were no detectable metabolic differences across the treatment groups.

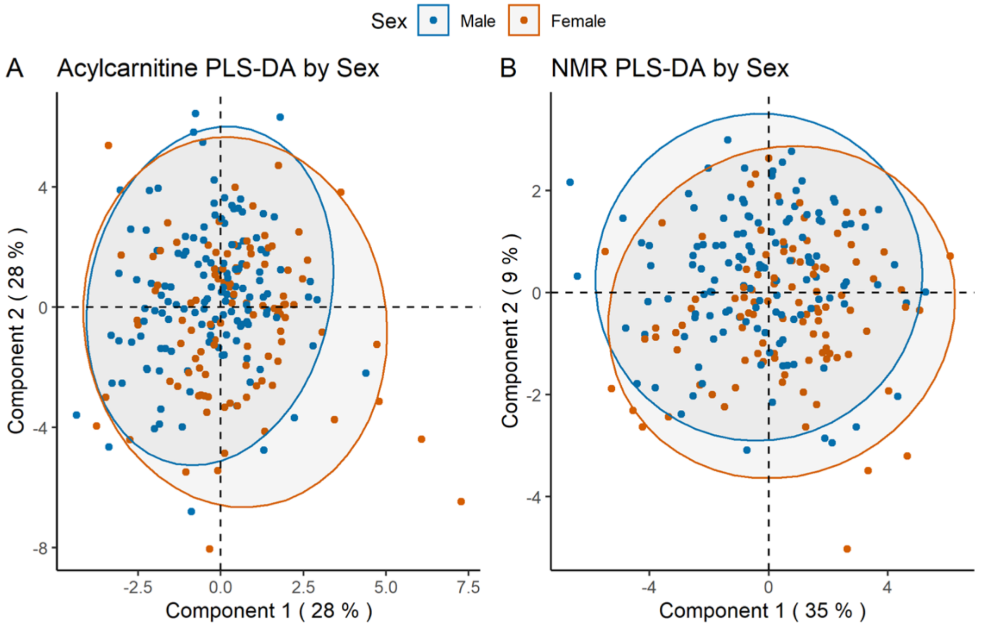


**Supplementary Figure 6**: Partial least squares-discriminant analysis (PLS-DA) does not differentiate acylcarnitines and nuclear magnetic resonance (NMR)-detected metabolites by sex in patients with septic shock. PLS-DA (and associated multivariate t distribution) of serum acylcarnitines (**A**) and the NMR-detected metabolites (**B**) from the 236 (102, female) and 228 (100, female) patients, respectively, of the 250 patients enrolled in the Rapid Administration of Carnitine (RACE) in Sepsis clinical trial.[1]

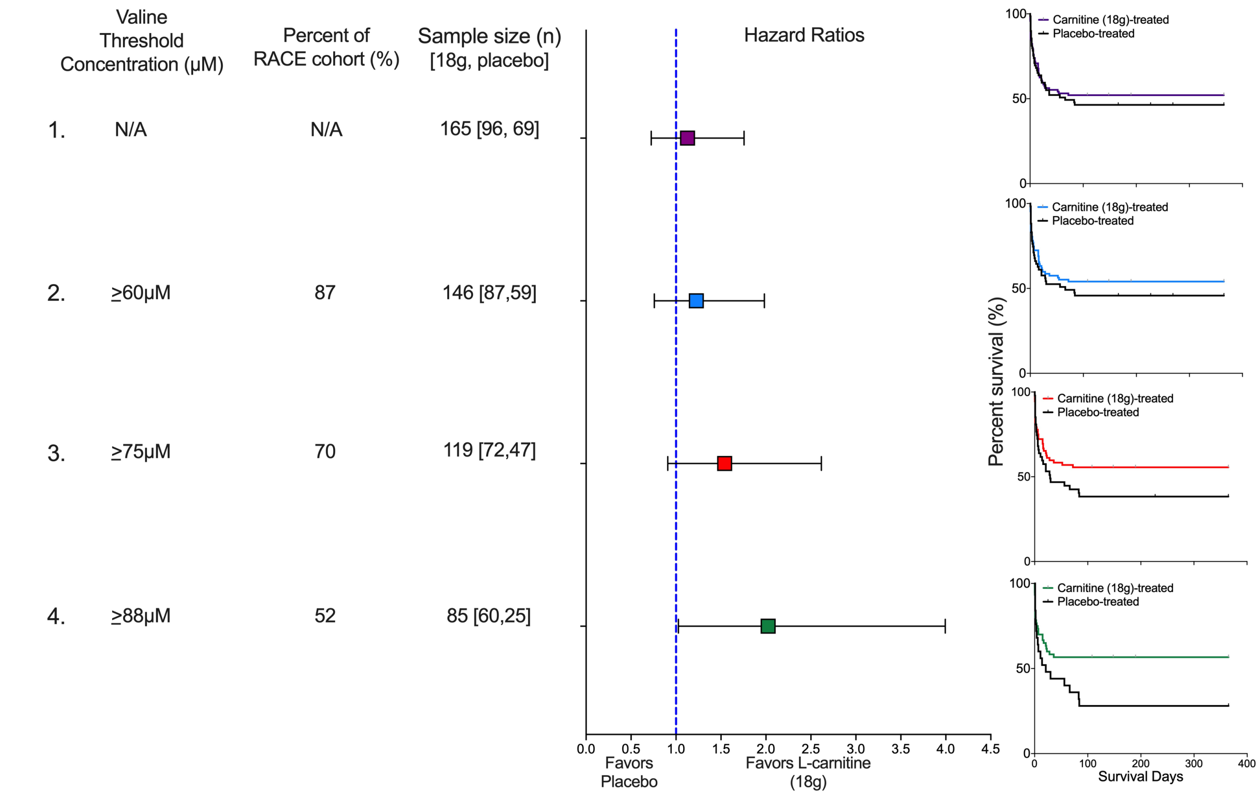


**Supplementary Figure 7. Pre-treatment valine concentration may be predictive of an L-carnitine treatment mortality benefit in patients with septic shock.** We present four scenarios to illustrate how different threshold concentrations of valine, a branched chain amino acid, would have impacted the outcome of the Rapid Administration of Carnitine (RACE) in Sepsis clinical trial in patients treated with either L-carnitine (18 g) or placebo. In scenario 1, no threshold concentration is used so the entire RACE cohort (n=228) is eligible. The sample size of 165 patients represents those that received either L-carnitine (18 g; n=96) or placebo (n=69). The hazard ratio is not significant, and consistent with the parent trial, the Kaplan-Meier curve shows no mortality benefit of L-carnitine (p=0·59). In scenario 2, a valine threshold concentration of ≥60µM is used. Eighty-seven percent (n=198) of the RACE cohort met this criterion and of these, 146 patients received either L-carnitine (18 g) or placebo. The hazard ratio is not improved, and the Kaplan-Meier curve shows no mortality benefit of L-carnitine (p=0·41). In scenario 3, valine threshold concentration of ≥75µM is used. Seventy percent (n=160) of the RACE cohort met this criterion and of these, 119 patients received either L-carnitine (18 g) or placebo. The hazard ratio is modestly improved but is not significant; the Kaplan-Meier curve shows no mortality benefit of L-carnitine (p=0·11). Finally, scenario 4 uses the valine concentration associated with the maximum Z-statistic (see Table 4 in the supplementary material), ≥88µM. Fifty-two percent (n=119) of the RACE cohort met this criterion and of these, 85 patients received either L-carnitine (18 g) or placebo. The hazard ratio is significant, and the Kaplan-Meier curve shows a mortality benefit of L-carnitine (p=0·04). Kaplan-Meier curves were interpreted using log-rank (Mantel-Cox) tests; hazard ratios were calculated using Mantel-Haenszel. The number of patients at risk at each time can be found here (DOIXX); no participants were censored.

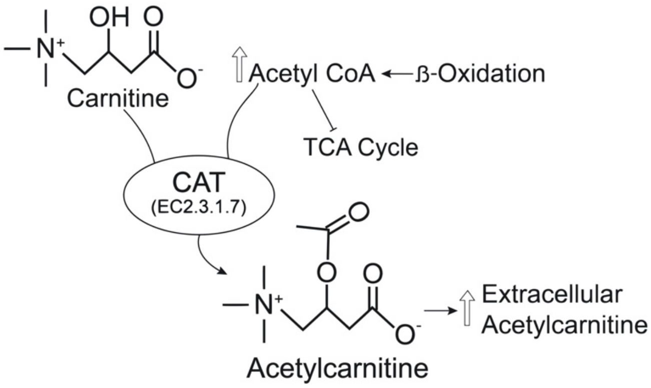


**Supplementary Figure 9**: In the mitochondria, sepsis induced altered flux through β-oxidation and TCA cycle energy pathways, including a possible TCA cycle “stall”, may lead to increased levels of acetyl-CoA. These excess acetyl-groups are removed via carnitine acetyl-transferase (CAT) mediated metabolism of carnitine converting it to acetylcarnitine (C2). In the setting of sepsis, this could be a plausible mechanism that explains the broad range of extracellular (serum) acetylcarnitine (C2) concentrations in sepsis patients. In patients with higher acetylcarnitine (C2), the mortality benefit of supplemental L-carnitine may be driven by it serving as a “sink” for excess acetyl-CoA/acetyl-groups.
